# Resource Profile: The Regenstrief Institute COVID-19 Research Data Commons (CoRDaCo)

**DOI:** 10.1101/2021.12.17.21267942

**Authors:** Katie S. Allen, Nader Zidan, Vishal Dey, Eneida A. Mendonca, Shaun Grannis, Suranga Kasturi, Babar Khan, Sarah Zappone, David Haggstrom, Laura Ruppert, Titus Schleyer, Xia Ning, Peter Embi, Umberto Tachinardi

## Abstract

The primary objective of the COVID-19 Research Data Commons (CoRDaCo) is to provide broad and efficient access to a large corpus of clinical data related to COVID-19 in Indiana, facilitating research and discovery. This curated collection of data elements provides information on a significant portion of COVID-19 positive patients in the State from the beginning of the pandemic, as well as two years of health information prior its onset. CoRDaCo combines data from multiple sources, including clinical data from a large, regional health information exchange, clinical data repositories of two health systems, and state laboratory reporting and vital records, as well as geographic-based social variables. Clinical data cover information such as healthcare encounters, vital measurements, laboratory orders and results, medications, diagnoses, the Charlson Comorbidity Index and Pediatric Early Warning Score, COVID-19 vaccinations, mechanical ventilation, restraint use, intensive care unit and ICU and hospital lengths of stay, and mortality. Interested researchers can visit ridata.org or to discuss access to CoRDaCo.

**Key Features:**

- CoRDaCo includes patient-level data on diagnosis and treatment, healthcare utilization, outcomes, and demographics. The level of detail available for each patient varies depending on the source of the clinical data.
- CoRDaCo uses geographic identifiers to link patient-specific data to area-level social factors, such as census variables and social deprivation indices.
- As of 4/30/21, the CoRDaCo cohort consists of over 776,000 cases, including granular data on over 15,000 patients who were admitted to an intensive care unit, and over 1,362,000 COVID-19-negative controls. Data is currently refreshed two times per month.
- The most prevalent comorbidities in the data set include hypertension, diabetes, chronic pulmonary disease, renal disease, cancer, and congestive heart failure.

## Data resource basics

The COVID-19 pandemic has presented the United States and the world with a challenge that requires rapid understanding of a novel and fast-moving infectious disease, for immediate treatment as well as analyzing long-term implications of the condition (1–5). To facilitate the research necessary to help understand this disease, comprehensive data is needed for all aspects of the condition, from disease onset to long-term complications. At the beginning of the crisis, clinicians and researchers needed to identify the clinical signs, symptoms, and characteristics, as well as the spectrum of trajectories of the disease, to effectively diagnose and manage patients. As the pandemic progressed, improved epidemiologic, diagnostic, and therapeutic approaches for managing long-term consequences became increasingly important with the emergence of a new condition - Post-Acute Sequelae of SARS-CoV-2 (PASC) Infection (Long Covid) (6). As more people are vaccinated, understanding the effectiveness of various vaccines, as well as the frequency and nature of reinfection, are becoming an added focus.

These characteristics of the pandemic highlight the need for data sources that are (1) comprehensive, i.e., by integrating data from multiple sources; (2) as complete as possible, i.e., by covering populations as thoroughly as possible; (3) extend, ideally, from the onset of the pandemic to the present; and (4) provide important context, such as preexisting conditions.

Many resources based on electronic health records (EHRs) and related data have been developed, including the National COVID Cohort Collaborative (N3C) (7–10). In mid-2020, the Regenstrief Institute (Regenstrief) leveraged its access to the Indiana Network for Patient Care (INPC) and selected health system data warehouses to create the COVID-19 Research Data Commons (CoRDaCo) to provide an efficient and effective way of generating timely datasets for COVID-19-related research. CoRDaCo combines multiple sources of Indiana-wide, EHR-derived clinical and “exposome” data (e.g., social determinants, mobility data, etc.), testing data, and vital statistics data (e.g., deaths) focused on COVID-19 case and control populations. A curated collection of data elements specific to COVID-19-positive patients, stored in a structured way, allows for more efficient creation of study-specific data sets. Additionally, creating this central repository allows for use of applications that will give researchers direct access to general data sets.

## Data sources

CoRDaCo leverages Regenstrief’s unique access to a vast Indiana-wide set of data from three types of sources: (1) the INPC, the state’s health information exchange; (2) COVID-19 testing and vaccination data from the Indiana State Department of Health (IDOH); and (3) data from the clinical data repositories of two major health systems in Indiana, Indiana University (IU) Health and Eskenazi Health:

1. <u>Indiana Network for Patient Care</u>. The INPC (11), managed by the Indiana Health Information Exchange (IHIE), is the oldest and one of the largest regional health information exchanges in the country. Established in the early 1990’s and expanded in 2004, it contains clinical elements from 123 separate healthcare entities, including major hospitals, health networks, insurance providers, state laboratory reporting, and the state vitals (death) reporting. Combined, the INPC contains data on over 18 million patients in the form of 15 billion clinical observations, and 319 million mineable text reports (12). While the INPC does not cover the entire State of Indiana, geographically and by population, it contains data on approximately 75% of the population, making this data source unique for its breadth of data coverage. Additionally, all patient addresses are geocoded and updated if they change, allowing for linkage to area-level social factors.
2. <u>Indiana State Department of Health</u>. IHIE and the IDOH have a long history of collaboration, including data sharing in support of public health needs such as emergency public health surveillance (13), and research access to Medicare and Medicaid claims data. The unprecedented need for public health data and research strengthened this relationship and includes near real-time data flow for laboratory reporting on COVID-19 testing and variants, vaccination data, and mortality data.
3. <u>Health System Data</u>. The clinical data repositories of two major health systems in Indiana, IU Health and Eskenazi Health, add valuable, granular data about patients’ healthcare events not contained in the INPC. With 18 hospitals and numerous outpatient facilities, IU Health has broad geographic coverage in Indiana. Eskenazi Health is a community safety-net system for Marion County, the largest county in the state, with one primary hospital and 11 outpatient facilities. While both systems contribute a significant portion of their data to the INPC, their clinical data repositories contain more detailed emergency department and inpatient records, enabling access to highly granular details related to patient care.

## Data elements

CoRDaCo integrates data from the above-mentioned clinical repositories to create a registry of patients with a clinical history of COVID-19 as well as COVID-19 negative controls in Indiana. The phenotype for identifying the COVID-19 positive patients in Indiana was created as part of Regenstrief’s partnership with IDOH during the COVID-19 mitigation efforts (14). This phenotype identifies all COVID-19 positive patients through laboratory testing and ICD code, relying primarily on state laboratory reporting, which encompasses all of Indiana. Controls are identified based upon presence of at least one negative COVID test and no positive COVID tests, and an INPC encounter in 2018 or 2019.

For each included patient – either as a case or control – CoRDaCo includes a minimum two-year look back period (to 1/1/2018) to identify pre-existing conditions, demographics, and limited medication fill data. A Charlson Comorbidity Index (15) is automatically calculated using the available data in this look-back period. For cases with a COVID-related hospitalization at either IU Health or Eskenazi Health – identified as an inpatient encounter 14 days prior to or following a positive COVID-19 test – additional granular details on the hospitalization are available. Community-level variables related to social deprivation index, tobacco access, air pollution, food insecurity, life expectancy, residential segregation, 211 calls for social services, and transit services are available for all cases and controls. Many of these variables are available through our partnership with the IUPUI Polis Center (16). Table 1 details the data elements included in CoRDaCo.

**Table 1.** details the general data elements included in CoRDaCo, with variability occurring between patients.

|  | Cases |  | Controls |
| --- | --- | --- | --- |
|  | No hospital stay | Hospital stay |  |
| Demographics | X | X | X |
| Clinical observations | X | X | X |
| Medication fill data | X | X | X |
| Mortality data | X | X | X |
| Health System Encounters | X | X | X |
| Diagnoses | X | X | X |
| Charlson Comorbidity Index | X | X | X |
| COVID-19 Vaccination data | X | X | X |
| Mechanical ventilation |  | X |  |
| Restraint use |  | X |  |
| ICU and hospital length of stay |  | X |  |
| Inpatient medications |  | X |  |
| Vital Measurements |  | X |  |
| Pediatric Early Warning Score (16) |  | X |  |
| Community-level variables | X | X | X |

Ongoing management of CoRDaCo’s phenotype and included data elements is overseen by a Steering Committee, which consists of clinical informatics experts as well as physicians representing infectious disease, intensive care, and pediatrics. Given the rapidly changing knowledge needs related to COVID-19 and PASC, an iterative process for phenotype implementation and data element inclusion was designed. The request for a change is analyzed by the data services team for consistency, integrability, completeness, and meaningfulness, and is channeled to the Steering Committee for discussion as needed. Changes to the data model are implemented in a systematic fashion on a routine schedule.

Future enhancements will include further utilization data for the time leading up to the pandemic, measurements indicating depth of data for individual patients, and conversion to the common data model OMOP to facilitate collaboration across organizations. Additionally, all current data in CoRDaCo are structured data. We are working on methods to extract valuable data contained in narrative text. The nDepth natural language processing system, developed by the Indiana Clinical and Translational Science Institute (CTSI) and Regenstrief, will be used to allow novel methods to identify essential findings in text reports, such as symptomology or family history, and further augment CoRDaCo (17–21).

## Data resource characteristics

National data suggest that Indiana is similar to US averages, increasing generalizability of studies produced from Indiana-derived datasets. In terms of high school graduation or higher (89% vs. 88%),(22,23) persons without health insurance (both 10%),(22,23) persons living in poverty (12% vs. 11%),(22,23) persons aged 65 and over (16% vs. 17%),(22,23) percentage female (both 51%),(22,23) percentage of low birthweight infants (both 8%),(24,25) and percentage of preterm infants (both 10%).(26,27) Indiana has a higher percentage of white race (85% vs. 76%),(22,23) and citizens living in rural locations (22% vs. 14%),(28) and a lower percentage of black race (10% vs. 13%)(22,23) than the US average.

Tables 2 and 3 below detail the baseline characteristics of the case and control patients. Continuous variables (e.g., “age”) are presented using median, minimum, maximum, mean, and standard deviation. Categorical variables (e.g., “gender”, “standard race”) are presented using counts and percentages. A patient was categorized as an ICU patient if s/he had an encounter record such that the ICU flag was set to ‘1’ and the admit time was after 3/6/2020. In addition, if a patient had an inpatient ICU encounter where the admit time was after 3/6/2020, s/he was also categorized as an ICU patient. All other patients were categorized as non-ICU.

**Table 2.**
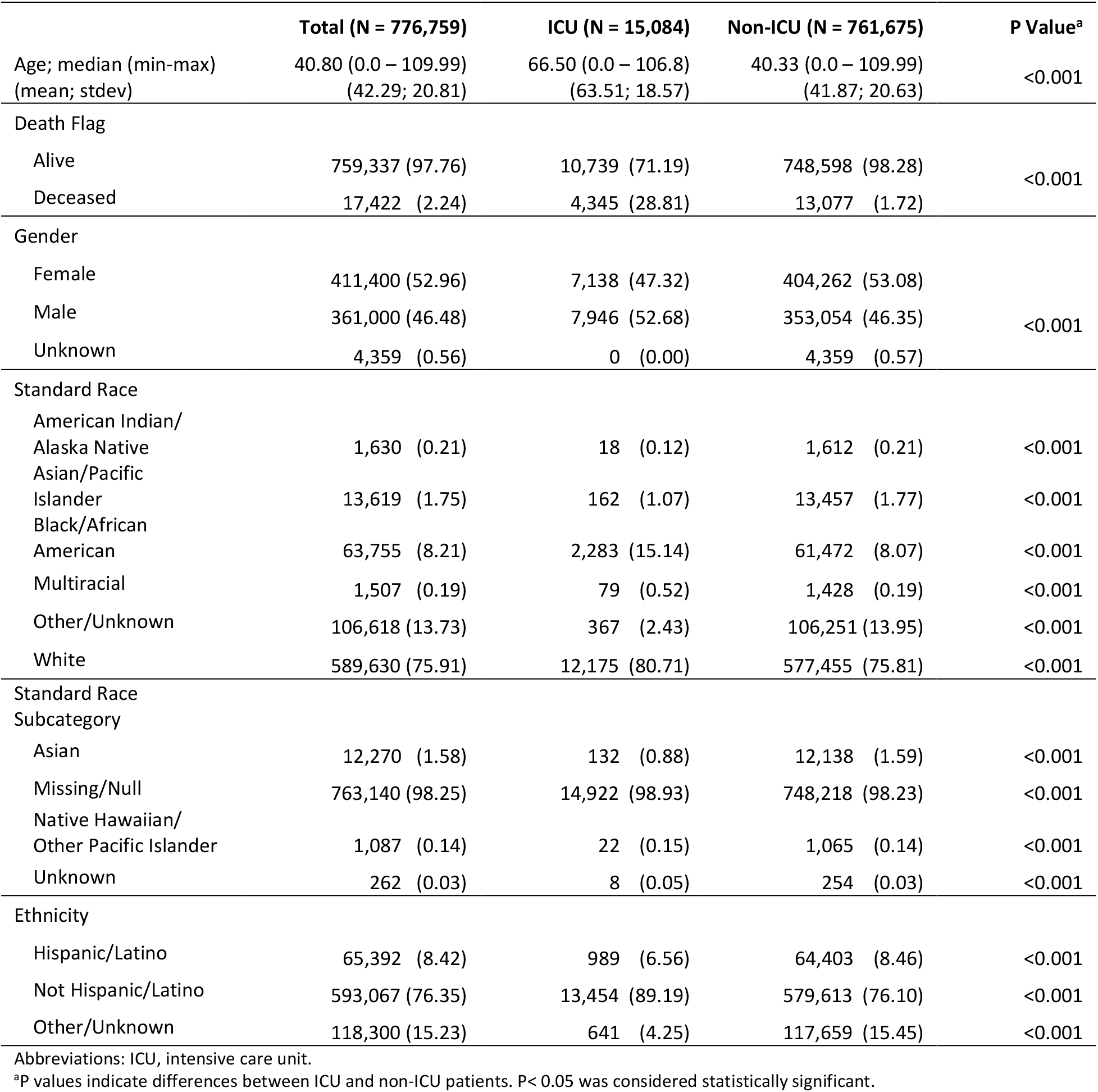
Baseline Characteristics of COVID-19-positive Patients

|  | **Total (N = 776,759)** | **ICU (N = 15,084)** | **Non-ICU (N = 761,675)** | **P Value<sup>a</sup>** |
| --- | --- | --- | --- | --- |
| Age; median (min-max)<br>(mean; stdev) | 40.80 (0.0 – 109.99)<br>(42.29; 20.81) | 66.50 (0.0 – 106.8)<br>(63.51; 18.57) | 40.33 (0.0 – 109.99)<br>(41.87; 20.63) | <0.001 |
| Death Flag |  |  |  |  |
| Alive | 759,337 (97.76) | 10,739 (71.19) | 748,598 (98.28) | <0.001 |
| Deceased | 17,422 (2.24) | 4,345 (28.81) | 13,077 (1.72) |  |
| Gender |  |  |  |  |
| Female | 411,400 (52.96) | 7,138 (47.32) | 404,262 (53.08) | <0.001 |
| Male | 361,000 (46.48) | 7,946 (52.68) | 353,054 (46.35) |  |
| Unknown | 4,359 (0.56) | 0 (0.00) | 4,359 (0.57) |  |
| Standard Race |  |  |  |  |
| American Indian/<br>Alaska Native | 1,630 (0.21) | 18 (0.12) | 1,612 (0.21) | <0.001 |
| Asian/Pacific<br>Islander | 13,619 (1.75) | 162 (1.07) | 13,457 (1.77) | <0.001 |
| Black/African<br>American | 63,755 (8.21) | 2,283 (15.14) | 61,472 (8.07) | <0.001 |
| Multiracial | 1,507 (0.19) | 79 (0.52) | 1,428 (0.19) | <0.001 |
| Other/Unknown | 106,618 (13.73) | 367 (2.43) | 106,251 (13.95) | <0.001 |
| White | 589,630 (75.91) | 12,175 (80.71) | 577,455 (75.81) | <0.001 |
| Standard Race<br>Subcategory |  |  |  |  |
| Asian | 12,270 (1.58) | 132 (0.88) | 12,138 (1.59) | <0.001 |
| Missing/Null | 763,140 (98.25) | 14,922 (98.93) | 748,218 (98.23) | <0.001 |
| Native Hawaiian/<br>Other Pacific Islander | 1,087 (0.14) | 22 (0.15) | 1,065 (0.14) | <0.001 |
| Unknown | 262 (0.03) | 8 (0.05) | 254 (0.03) | <0.001 |
| Ethnicity |  |  |  |  |
| Hispanic/Latino | 65,392 (8.42) | 989 (6.56) | 64,403 (8.46) | <0.001 |
| Not Hispanic/Latino | 593,067 (76.35) | 13,454 (89.19) | 579,613 (76.10) | <0.001 |
| Other/Unknown | 118,300 (15.23) | 641 (4.25) | 117,659 (15.45) | <0.001 |

Abbreviations: ICU, intensive care unit.
<sup>a</sup>P values indicate differences between ICU and non-ICU patients. P< 0.05 was considered statistically significant.

**Table 3.**
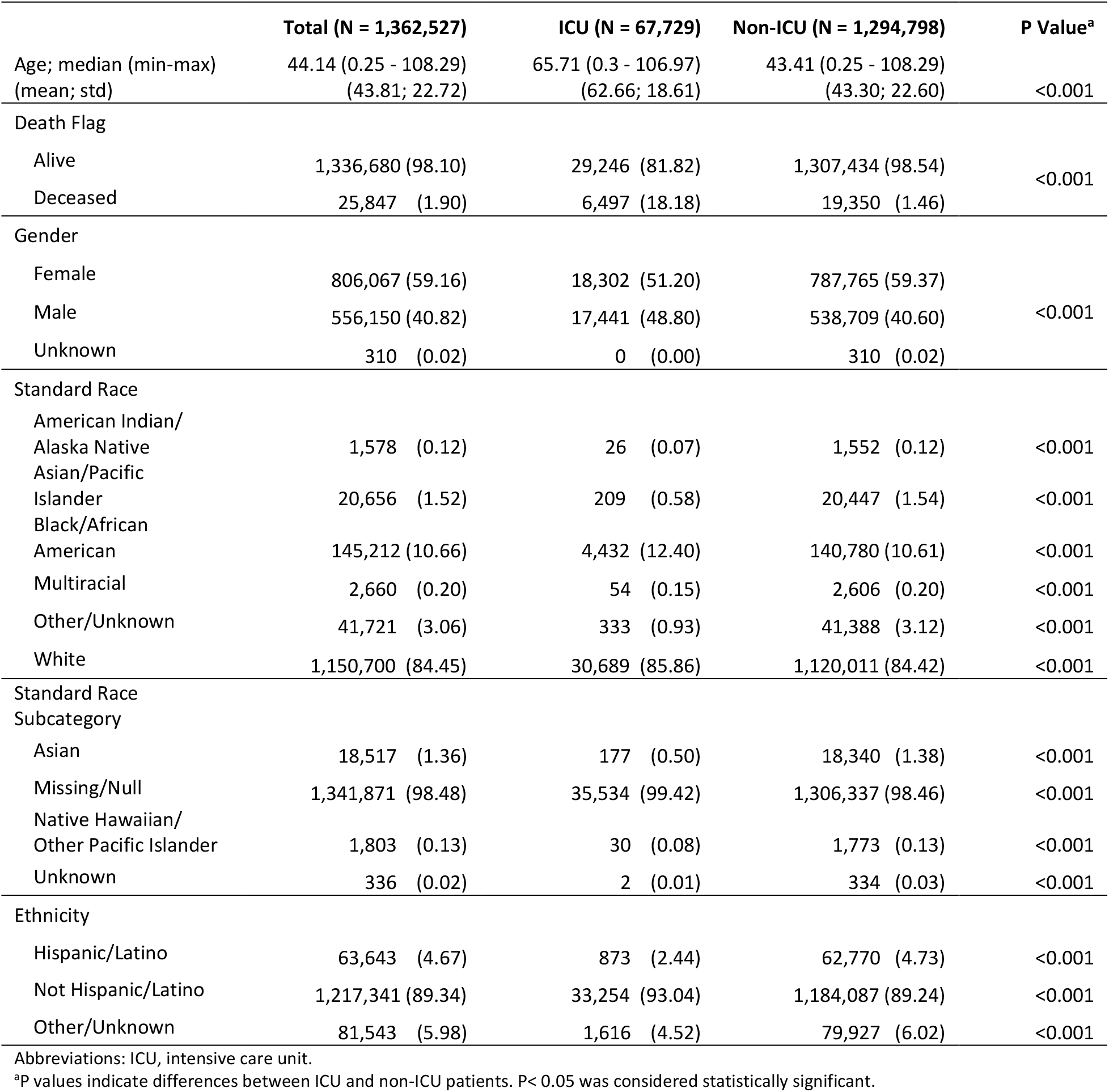
Baseline Characteristics of non-COVID-19 Control Patients

|  | **Total (N = 1,362,527)** | **ICU (N = 67,729)** | **Non-ICU (N = 1,294,798)** | **P Value<sup>a</sup>** |
| --- | --- | --- | --- | --- |
| Age; median (min-max)<br>(mean; std) | 44.14 (0.25 - 108.29)<br>(43.81; 22.72) | 65.71 (0.3 - 106.97)<br>(62.66; 18.61) | 43.41 (0.25 - 108.29)<br>(43.30; 22.60) | <0.001 |
| Death Flag |  |  |  |  |
| Alive | 1,336,680 (98.10) | 29,246 (81.82) | 1,307,434 (98.54) | <0.001 |
| Deceased | 25,847 (1.90) | 6,497 (18.18) | 19,350 (1.46) |  |
| Gender |  |  |  |  |
| Female | 806,067 (59.16) | 18,302 (51.20) | 787,765 (59.37) | <0.001 |
| Male | 556,150 (40.82) | 17,441 (48.80) | 538,709 (40.60) |  |
| Unknown | 310 (0.02) | 0 (0.00) | 310 (0.02) |  |
| Standard Race |  |  |  |  |
| American Indian/<br>Alaska Native | 1,578 (0.12) | 26 (0.07) | 1,552 (0.12) | <0.001 |
| Asian/Pacific<br>Islander | 20,656 (1.52) | 209 (0.58) | 20,447 (1.54) | <0.001 |
| Black/African<br>American | 145,212 (10.66) | 4,432 (12.40) | 140,780 (10.61) | <0.001 |
| Multiracial | 2,660 (0.20) | 54 (0.15) | 2,606 (0.20) | <0.001 |
| Other/Unknown | 41,721 (3.06) | 333 (0.93) | 41,388 (3.12) | <0.001 |
| White | 1,150,700 (84.45) | 30,689 (85.86) | 1,120,011 (84.42) | <0.001 |
| Standard Race<br>Subcategory |  |  |  |  |
| Asian | 18,517 (1.36) | 177 (0.50) | 18,340 (1.38) | <0.001 |
| Missing/Null | 1,341,871 (98.48) | 35,534 (99.42) | 1,306,337 (98.46) | <0.001 |
| Native Hawaiian/<br>Other Pacific Islander | 1,803 (0.13) | 30 (0.08) | 1,773 (0.13) | <0.001 |
| Unknown | 336 (0.02) | 2 (0.01) | 334 (0.03) | <0.001 |
| Ethnicity |  |  |  |  |
| Hispanic/Latino | 63,643 (4.67) | 873 (2.44) | 62,770 (4.73) | <0.001 |
| Not Hispanic/Latino | 1,217,341 (89.34) | 33,254 (93.04) | 1,184,087 (89.24) | <0.001 |
| Other/Unknown | 81,543 (5.98) | 1,616 (4.52) | 79,927 (6.02) | <0.001 |

Abbreviations: ICU, intensive care unit.
<sup>a</sup>P values indicate differences between ICU and non-ICU patients. P< 0.05 was considered statistically significant.

Table 2 presents the baseline demographic characteristics of all COVID-19 patients diagnosed between 1/1/2020 and 4/30/21 in the entire CoRDaCo dataset. After removing patients with an age less than 0, greater than 110, or no age reported (which were likely to be errors during data entry), there were 776,759 patients in total diagnosed with COVID-19 with a median age of 40.8 years and a mean age of 42.3 years (interquartile range: 25.0 – 57.6). Among the 776,759 patients, 15,084 (1.94%) were admitted to the ICU with a median age of 66.50 years and a mean age of 63.5 years. In terms of gender, females constituted more than half of the COVID-19 patients (52.96%), slightly above from the rate in the U.S. population (51%)(23). Among ICU admitted patients, females constituted less than half (47.3%); however, among the patients not admitted to the ICU, females constituted more than half again (53.1%).

Table 3 presents the demographic characteristics of 1,362,527 COVID-19-negative control patients. The healthy controls are selected per the N3C phenotype, essentially representing individuals with at least one negative COVID-19 lab test and no positive tests during from 1/1/2020 - present. Additionally, these patients were required to have a clinical encounter between 2018-2019 to eliminate patients for whom the sole clinical data element is a COVID-19 test result. Note that the COVID-19-negative controls were not diagnosed with COVID-19 but may have other diseases.

Figure 1 shows the number and monthly percentage of clinical data elements in CoRDaCo by category from 3/1/2020 to 4/30/2021 in correlation with the number of COVID-19-positive cases in Indiana. For each category, we first calculated the total the number of records for the entire period, then the percentage for each month (indicated by hue and saturation). For encounters, the admission time was used for assignment to a specific month. To contextualize the monthly percentages, the top of Figure 1 shows the COVID-19 positivity trends for Indiana.

**Figure 1.**
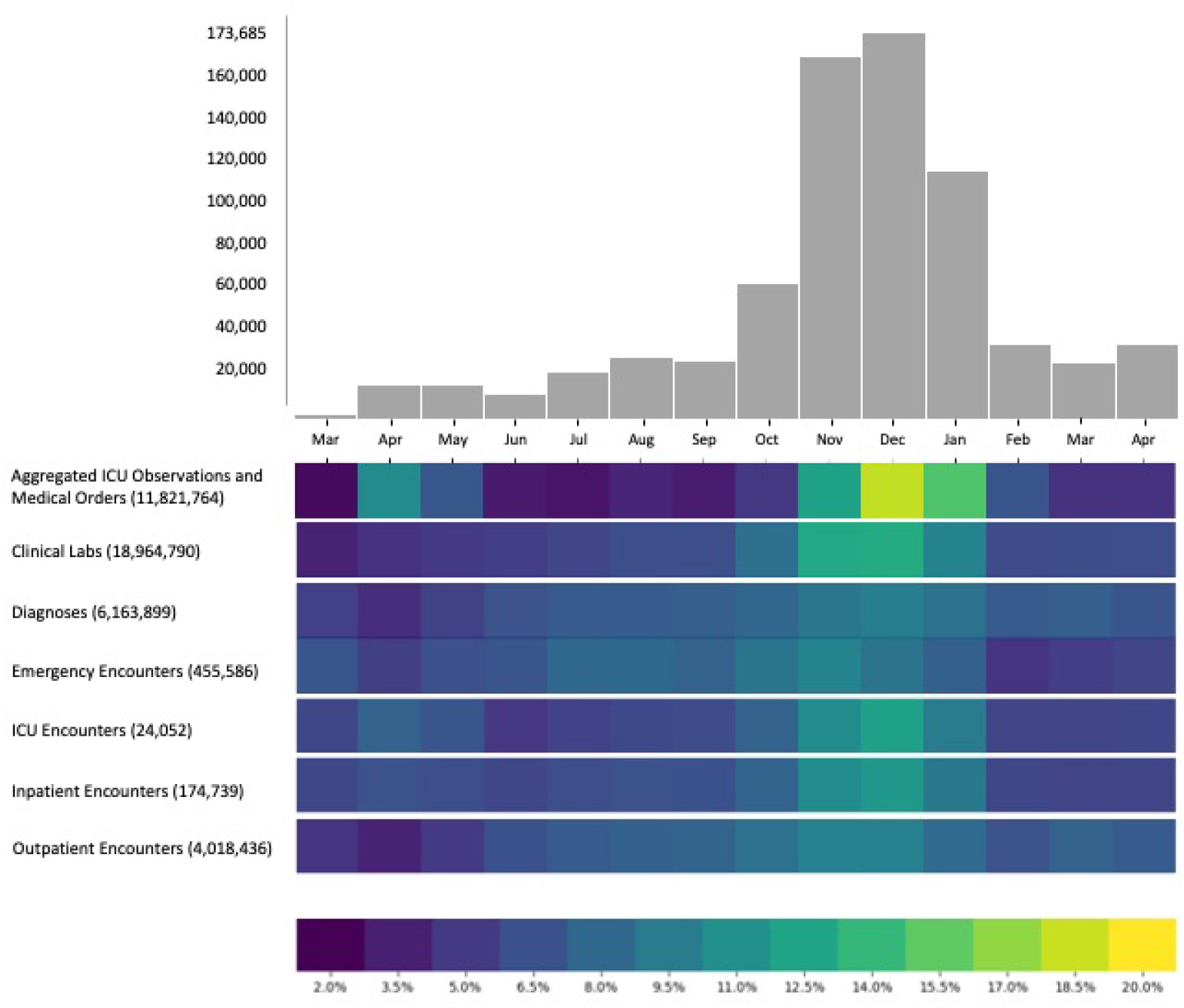
depicts COVID-19 positive cases by month from March 2020 to April 2021 with relative distribution of the number of records in each information category; total number of records in parentheses.

Figure 1 shows two general trends. First, the amount of data increases from early 2020 and reaches a peak in Nov./Dec. of 2020, followed by a decrease. While not empirically tested, this aligns with Indiana’s COVID-19 surge during the latter half of 2020, followed by a reduction in overall cases following the first of the year. April 2020 shows a decrease, specifically in diagnoses, and emergency and outpatient encounters. Again, while not empirically tested, this aligns with the Indiana Government Executive Order implementing shelter-in-place and reduced services throughout the State, including reductions in non-emergency care (29).

Table 4 shows the total number of unique patients with data by category and year. The number of patients shows an increasing trend in most categories, except for “inpatient only” and “outpatient only” categories. This exception may be related to the presence of more types of data in the system, either due to increased healthcare utilization or the increase in health system participation during the pandemic. Of note, for 18.4% of the cohort, we only have the positive COVID-19 test results and no other data elements.

**Table 4.**
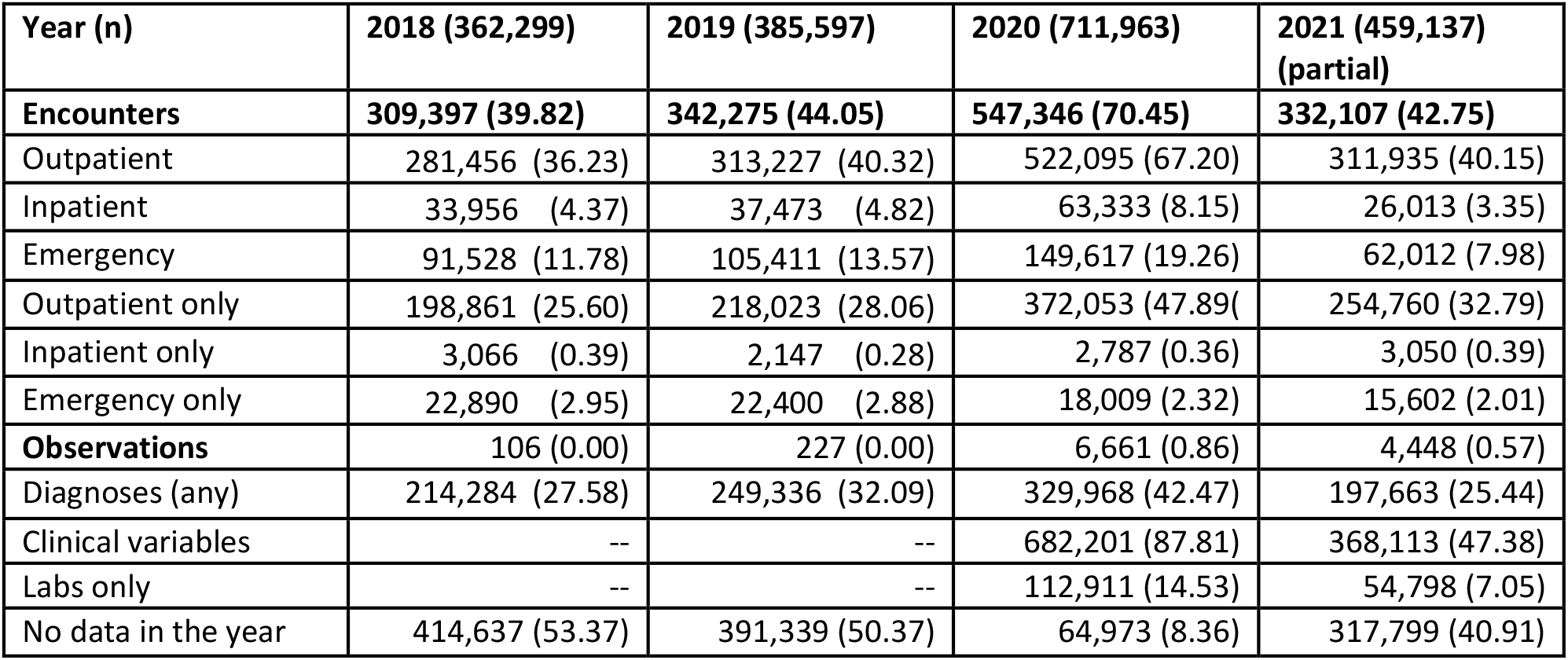
Total number of unique patients with data by category and year. Percentages in parentheses (except Row 1).

CoRDaCo may also be suitable for comorbidity-specific studies. Table 5 shows the number of patients in the case and control cohort with comorbid conditions associated with COVID-19 complications (30).

**Table 5.** Number of COVID-19-positive and -negative patients with specific comorbidity

| **Comorbidity** | **Patient count (Percentage)** |  |
| --- | --- | --- |
|  | **COVID-19-positive** | **COVID-19-negative** |
| Hypertension | 91,441 (11.77%) | 306,758 (22.51%) |
| Diabetes | 53,865 (7%) | 163,489 (12%) |
| Chronic Pulmonary Disease | 41,901 (5.39%) | 153,722 (11.28%) |
| Renal Disease | 17,617 (2.27%) | 58,833 (4.32%) |
| Cancer | 17,139 (2.21%) | 75,225 (5.52%) |
| Congestive Heart Failure | 16,798 (2.16%) | 60,266 (4.42%) |
| Cerebrovascular Disease | 11,951 (1.54%) | 45,928 (3.37%) |
| Myocardial Infarction | 7,500 (0.97%) | 27,968 (2.05%) |
| Moderate or Severe Liver Disease | 604 (0.08%) | 2,924 (0.21%) |

## Strengths and weaknesses

CoRDaCo data are a valuable source of detailed longitudinal information on COVID-19 patients in Indiana. It is important that users understand the complexities of using these predominantly clinical data for research purposes to help them develop accurate interpretations. Almost all data in the CoRDaCo are real-world, electronic health record data recorded by clinicians in many healthcare organizations and practice settings. Differences in organizational culture, clinician preferences, limited standards for data collection, IT system limitations and many other factors cause variations in the data. Most of the limitations mentioned below are implicit in EHR data and not necessarily unique to CoRDaCo.

### Strengths

A significant strength of CoRDaCo is its size, level of detail, and geographic coverage. Our partnership with the IDOH ensures that CoRDaCo contains all COVID-19 test results within the state. CoRDaCo links these results to a significant proportion of the healthcare information representing 75% of Indiana residents. This allows us to put the COVID-19 status of each person in the context of their overall healthcare experience and utilization. Since CoRDaCo is a real-time registry that is fed by operational systems, data accumulate continually and can help tell the longitudinal patient story across large numbers of individuals. The transmission delays common in maintaining many registries automatically are not an issue for CoRDaCo since clinical data are transferred to our research databases within a few minutes of being generated. In addition, data from different sources about the same patient are automatically aggregated using state-of-the-art patient matching algorithms. Using EHR data reduces subjective biases found in self-reported health surveys, since EHR data comprise professionally generated diagnoses, laboratory and examination results, and prescriptions. The broad representation of a demographically and geographically diverse patient population that resembles the US population at large makes CoRDaCo suitable for population-health level analytics and decision-making.

### Challenges and weaknesses

As mentioned above, one of the biggest challenges is that the data that constitute CoRDaCo are generated through clinical practice and public health, not research. Thus, CoRDaCo data do not have the high degree of standardization, homogeneity, and quality that data generated and properly curated in high-quality research studies have. The characteristics of the INPC, a major data source for CoRDaCo, cause certain biases. As previously mentioned, not all healthcare organizations in Indiana transmit data to the INPC. Represented healthcare organizations are predominately urban. Additionally, organizations contributing data may only send a selection of data deemed to be most relevant to general clinical care. This leaves a gap with relation to certain data elements, such as medications. This is somewhat mitigated by the augmentation of INPC data with assess to the warehouses for two major health systems, however this is only for a subset of patients.

## Data resource access

CoRDaCo is currently accessible through three mechanisms, two mediated by the Regenstrief Data Services (RDS) team and the third that is accessible with fewer restrictions.

1. **Custom data exports by RDS data managers (mediated)**: This method of data access produces custom data sets that can either be completely de-identified or contain some identifiable information.
2. **Research data networks**: Research data networks produce larger study cohorts by combining data from various sites. Regenstrief participates in the N3C, the Chicago Area Patient-Centered Outcomes Research Network (CAPriCORN), and CTSI Accrual to Clinical Trials (ACT) Network. We contribute selected CoRDaCo data to these initiatives. Researchers can access CoRDaCo data in accordance with initiative-specific protocols.
3. **User exploration and synthetic CoRDaCo data sets**: A front facing query tool (MDClone ©) is available to allow users to explore the data available in CoRDaCo. The user can create queries to explore the feasibility of use cases. The queries can be shared with Regenstrief data analysts for more efficient data set creation. Alternatively, these queries can be used to generate computationally derived synthetic data sets do not share mutual information with source data, eliminating re-identification potential. Synthetic CoRDaCo data sets will provide quick and efficient access to those corpora of data without much of the overhead involved in the preceding methods.

Access to this data is available with proper governance in place. To request a customized data set, please visit ridata.org and complete the request form. Direct access to CoRDaCo data is also available via MDClone © accounts for which are available at no-cost. To request access to this tool, visit ridata.org and complete the account request form

for that or any other questions regarding CoRDaCo.

## Data Availability

Access to this data is available with proper governance in place. To request a customized data set, please visit ridata.org and complete the request form. Direct access to CoRDaCo data is also available via MDClone accounts for which are available at no-cost. To request access to this tool, visit ridata.org and complete the account request form
for that or any other questions regarding CoRDaCo.

## Ethics approval statement

This data resource was created with approval from the Indiana University Institutional Review Board, protocol number 12712.

## Acknowledgements

The authors would like to acknowledge the work of the dedicated data analysts who brought CoRDaCo to fruition, including Lauren Lembcke, John Price, Amy Hancock, and Jack Vanschaik. We would also like to acknowledge the non-authoring members of the CoRDaCo Steering Committee meetings: Sikander Khan, Haley Pritchard, and Thankam Thyvalikakath. This project was made possible, in part, by support from the National Library Of Medicine of the National Institutes of Health under Award Number R01LM012605, the Regenstrief Institute funding to support research projects focused on SARS-CoV-2 (COVID-19), the Indiana Clinical and Translational Sciences Institute (funded in part by Award Number UL1TR002529 from the National Institutes of Health, National Center for Advancing Translational Sciences) Clinical and Translational Sciences Award, and the Lilly Endowment, Inc. Physician Scientist Initiative. Any opinions, findings, and conclusions or recommendations expressed in this material are those of the authors, and do not necessarily reflect the views of the funding agencies.

